# COVID-19 is associated with multiple sclerosis exacerbations that are prevented by disease modifying therapies

**DOI:** 10.1101/2021.03.08.21253141

**Authors:** Afagh Garjani, Rodden M Middleton, Rachael Hunter, Katherine A Tuite-Dalton, Alasdair Coles, Ruth Dobson, Martin Duddy, Stella Hughes, Owen R Pearson, David Rog, Emma C Tallantyre, Roshan das Nair, Richard Nicholas, Nikos Evangelou

## Abstract

**Background:** Infections can trigger exacerbations of multiple sclerosis (MS). The effects of the coronavirus disease 2019 (COVID-19) on MS are not known. The aim of this study was to understand the impact of COVID-19 on new and pre-existing symptoms of MS.

**Methods:** The COVID-19 and MS study is an ongoing community-based, prospective cohort study conducted as part of the United Kingdom MS Register. People with MS and COVID-19 were invited by email to complete a questionnaire about their MS symptoms during the infection. An MS exacerbation was defined as developing new MS symptoms and/or worsening of pre-existing MS symptoms.

**Results:** Fifty-seven percent (230/404) of participants had an MS exacerbation during their infection; 82 developed new MS symptoms, 207 experienced worsened pre-existing MS symptoms, and 59 reported both. Disease modifying therapies (DMTs) reduced the likelihood of developing new MS symptoms during the infection (OR 0.556, 95%CI 0.316-0.978). Participants with a higher pre-COVID-19 webEDSS (web-based Expanded Disability Status Scale) score (OR 1.251, 95%CI 1.060-1.478) and longer MS duration (OR 1.042, 95%CI 1.009-1.076) were more likely to experience worsening of their pre-existing MS symptoms during the infection.

**Conclusion:** COVID-19 infection was associated with exacerbation of MS. DMTs reduced the chance of developing new MS symptoms during the infection.

## INTRODUCTION

The role of systemic infections in provoking exacerbations of multiple sclerosis (MS) is well described.^1^ The coronavirus disease 2019 (COVID-19) is a viral infection, the effects of which on MS exacerbations have not been established. Understanding the impact of COVID-19 on MS symptoms will allow for thorough counselling of people with MS regarding the risk of infection during periods of community transmission.

Potential safety concerns about using immunosuppressive MS disease modifying therapies (DMTs) during the COVID-19 pandemic,^2^ along with disruptions to MS services,^3^ have resulted in changes to the treatment plans of many people with MS. However, a decrease in the use of DMTs during the pandemic could lead to excessive MS relapses. Further understanding of the relationship between COVID-19, MS relapses and DMTs will inform decision-making about altering or delaying treatment with DMTs.

In this paper, we study the impact of COVID-19 on pre-existing and new symptoms of MS in a large cohort of people with MS and COVID-19. We also assess potential factors associated with COVID-19 related MS exacerbations.

## MATERIALS AND METHODS

The COVID-19 and MS study is an ongoing national community-based, prospective cohort study conducted as part of the United Kingdom (UK) MS Register (UKMSR) ^4^. People with MS report whether they have had symptoms consistent with COVID-19, whether the diagnosis was confirmed by a healthcare provider based on their clinical or laboratory findings, and whether they have been admitted to a hospital because of their infection ^4^.

People with MS and symptoms consistent with COVID-19 were invited to complete a questionnaire about their MS symptoms during or soon after the infection between 20th of July 2020 and 25th of January 2021. We asked participants about any new or worsened pre-existing MS symptoms. Here, we report our cross-sectional findings according to the STROBE guidelines^5^.

We defined an MS exacerbation as developing new MS symptoms, worsening of pre-existing MS symptoms, or experiencing both during a COVID-19 infection. We asked participants about limitation in daily activities caused by the new symptoms and classified them as mild (no limitation), moderate (less than 50% limitation), or severe (more than 50% limitation).

We correlated the COVID-19 and MS symptoms data with information held by the UKMSR on participants’ demographics (age, sex, and ethnicity), clinical characteristics (MS type, disease duration from diagnosis, and DMTs), most recent recorded web-based Expanded Disability Status Scale (webEDSS) scores (scored 0 – 10, with higher scores indicating more neurological impairment) from before their infection ^6^, and most recent Hospital Anxiety and Depression Scale scores (scored 0 – 21, with scores ≥11 considered as probable cases of anxiety or depression) ^7^.

### Statistical Analysis

Data were analysed using IBM SPSS Statistics for Windows, version 26 (IBM Corp., Armonk, N.Y., USA; 2019).

Continuous data were compared using the independent samples t-test, if normally distributed (mean [standard deviation, SD]) or the Mann-Whitney U test, when not normally distributed (also used for comparing ordinal variables; median [interquartile range, IQR]). Categorical variables were analysed using the Chi-square test (or Fischer’s exact test if expected count ≤5). For variables with missing data, the number of valid values is stated.

The association between different dependent (developing new MS symptoms, worsening of pre-existing MS symptoms) and independent variables (age, sex, type of MS, MS disease duration, pre-COVID-19 webEDSS score, DMT use) was assessed using univariable or multivariable binomial logistic regression analysis. To avoid introducing bias by controlling for colliders and mediators in the regression analyses models, directed acyclic graphs (DAGs) were built to determine confounding factors for individual regression analyses ^89^.

Confounding factors controlled for in each analysis have been stated. Listwise deletion was implemented for missing data. The results of the regression analyses are presented as odds ratio (OR) and 95% confidence intervals (95% CI).

### Standard Protocol Approvals, Registrations, and Patient Consents

Ethical approval for UKMSR studies was obtained from South West-Central Bristol Research Ethics Committee (16/SW/0194). Participants provided informed consent online. The study is registered on clinicaltrials.gov: NCT04354519.

### Data Availability Policy

Data are stored on the UKMSR Secure e-Research Platform at Swansea University Medical School. Line level data cannot be released, but qualified researchers, subject to governance, can request access to data.

## RESULTS

We invited 978 people with MS and COVID-19 to complete the MS symptoms questionnaire and 404 (41%) responded within a median (IQR) duration of 14 (9 – 17) weeks from reporting a diagnosis of COVID-19 (Table 1).

**Table 1.** Demographic and clinical characteristics of participants and non-participants

|  | Participants<br>(n =404) | Non-participants<br>(n =574) | *p* value |
| --- | --- | --- | --- |
| Age, years, mean (SD) | 50 (11) | 48 (11) | 0.001 |
| Female, n (%) | 307 (76) | 456 (79.4) | 0.434 |
| White ethnicity, n (%) | 380 (94.1) | 538 (93.7) | 0.832 |
| Pre-COVID-19 webEDSS score <sup>a</sup>,<br>median (IQR) | 4.5 (3 – 6.5)<br>n =248 | 4 (3 – 6.5)<br>n =288 | 0.776 |
| MS type, n (%) |  |  |  |
| RRMS | 277 (68.6) | 415 (72.3) | 0.018 |
| SPMS | 65 (16.1) | 99 (17.2) |  |
| PPMS | 39 (9.7) | 26 (4.5) |  |
| Unknown | 23 (5.7) | 34 (5.9) |  |
| MS disease duration, years, median<br>(IQR) | 11 (5 – 18)<br>n =395 | 10 (5 – 17)<br>n =547 | 0.106 |
| DMTs, n (%) | 193 (47.8) | 301 (52.4) | 0.151 |
| Beta interferons | 21 (5.2) | 39 (6.8) |  |
| Glatiramer acetate | 22 (5.4) | 37 (6.5) |  |
| Teriflunomide | 7 (1.7) | 14 (2.4) |  |
| Dimethyl fumarate | 58 (14.4) | 72 (12.6) |  |
| Fingolimod | 24 (5.9) | 35 (6.1) |  |
| Siponimod | 0 (0) | 1 (0.2) |  |
| Natalizumab | 24 (5.9) | 44 (7.7) |  |
| Ocrelizumab | 14 (3.5) | 33 (5.8) |  |
| Cladribine | 7 (1.7) | 9 (1.6) |  |
| Alemtuzumab | 13 (3.2) | 15 (2.6) |  |
| Others <sup>b</sup> | 3 (0.7) | 1 (0.2) |  |
| Confirmed COVID-19, n (%) | 108 (26.7) | 168 (29.3) | 0.386 |
| Hospitalized due to COVID-19, n (%) | 8 (2) | 9 (1.6) | 0.620 |
| DMTs = Disease Modifying Therapies; IQR = Interquartile Range; PPMS = Primary Progressive MS; RRMS = Relapsing Remitting MS; SD = Standard Deviation; SPMS = Secondary Progressive MS; webEDSS = web-based Expanded Disability Status Scale<br><sup>a</sup> The median (IQR) duration from recording the webEDSS score to reporting COVID-19 was 7 (3 – 16.75) weeks for participants and 11 (6 – 23.75) weeks for non-participants ($p < 0.001$).<br><sup>b</sup> Participants were taking Ponesimod (n =1) and Rituximab (n =2) and the non-participant was taking Azathioprine. | | | |

Two hundred and thirty (57%) participants had an MS exacerbation, with 82 (20%) developing new symptoms, 207 (51%) experiencing worsened pre-existing symptoms, and 59 (15%) reporting both during their COVID-19 infection.

Ninety-seven percent (n =222) of participants with an MS exacerbation (80 with new MS symptoms and 199 with worsened pre-exiting MS symptoms) had fever during their infection compared to 68% (n =72) of participants without an MS exacerbation (*p* <0.001). Six (3%) participants with an MS exacerbation (2 with new MS symptoms and all 6 with worsened pre-existing MS symptoms) and 2 (1%) participants without an MS exacerbation were hospitalized due to COVID-19 (*p* =0.296).

The rate of MS exacerbations was not significantly different between participants with (n =108) and without (n =296) a confirmed diagnosis of COVID-19 (63.9% vs 54.4%, *p* =0.088).

A higher proportion of participants with anxiety and/or depression reported an MS exacerbation during their infection compared to participants without anxiety or depression (68% [78/114] vs 51% [109/212], *p* =0.003), with 32% (n =36) and 14% (n =30) reporting new MS symptoms, respectively, and 61% (n =69) and 48% (n =101) reporting worsened pre-existing MS symptoms, respectively.

Thirty-nine percent (77/196) of the participants with an MS exacerbation required additional support for their daily activities during COVID-19 infection, as opposed to only 6% (7/114) of the participants without an exacerbation (p <0.001).

### New MS Symptoms

Among the 82 participants with new MS symptoms during the infection, the most reported new symptoms were sensory, motor, or both (n =58; 71%) (Table 2). Some COVID-19 symptoms such as fatigue, memory problems, or mobility problems can mimic MS symptoms. Most participant who reported fatigue (n =18), memory problems (n =17), or mobility problems (n =24) as part of their new MS symptoms during the infection had additional non-COVID-19 related neurological symptoms including sensory, motor, visual, or balance problems (89%, 88%, and 71%, respectively).

**Table 2.** Reported new multiple sclerosis symptoms during COVID-19 infection

|  | N (%) |
| --- | --- |
| Symptoms <sup>a</sup> | 82 (100) |
| Weakness | 27 (6.7) |
| Mild | 6 (22.2) |
| Moderate | 14 (51.9) |
| Severe | 7 (25.9) |
| Sensory symptoms (numbness, pins and needles, or pain) | 43 (10.6) |
| Mild | 12 (30.8) |
| Moderate | 24 (61.5) |
| Severe | 7 (17.9) |
| Balance problems | 24 (5.9) |
| Mild | 5 (20.8) |
| Moderate | 14 (58.3) |
| Severe | 5 (20.8) |
| Bladder or bowel problems | 15 (3.7) |
| Mild | 4 (28.6) |
| Moderate | 6 (42.9) |
| Severe | 5 (35.7) |
| Visual problems (blurred vision or double vision) | 12 (3) |
| Mild | 5 (41.7) |
| Moderate | 3 (25) |
| Severe | 4 (33.3) |
| Fatigue | 18 (4.5) |
| Mild | 3 (16.7) |
| Moderate | 6 (33.3) |
| Severe | 9 (50) |
| Memory problems | 17 (4.2) |
| Mild | 3 (17.6) |
| Moderate | 9 (52.9) |
| Severe | 5 (29.4) |
| Mobility problems | 24 (5.9) |
| Mild | 3 (12.5) |
| Moderate | 13 (54.2) |
| Severe | 8 (33.3) |
| Others <sup>b</sup> | 10 (2.5) |
| <sup>a</sup> Symptoms causing no limitation in daily activities were considered as mild, symptoms causing less than 50% limitation in daily activities as moderate, and symptoms causing more than 50% limitation in daily activities as severe.<br><sup>b</sup> Other new MS symptoms included spasms, speech or swallowing difficulties, tremor, or vertigo |  |

Sixteen (20%) participants with new MS symptoms during their infection had mild, 40 (49%) had moderate, and 26 (32%) had severe symptoms. None were treated with steroids.

Taking DMTs reduced the likelihood of developing new MS symptoms during the infection (adjusted OR 0.556, 95% CI 0.316 – 0.978) (Table 3). We did not formally test the association between individual DMTs and developing new MS symptoms due to the small number of participants on individual DMTs; however, it seemed that a higher proportion of participants without new MS symptoms during their infection were taking fingolimod, ocrelizumab, or cladribine compared to participants who developed new symptoms (Table 4).

**Table 3.**
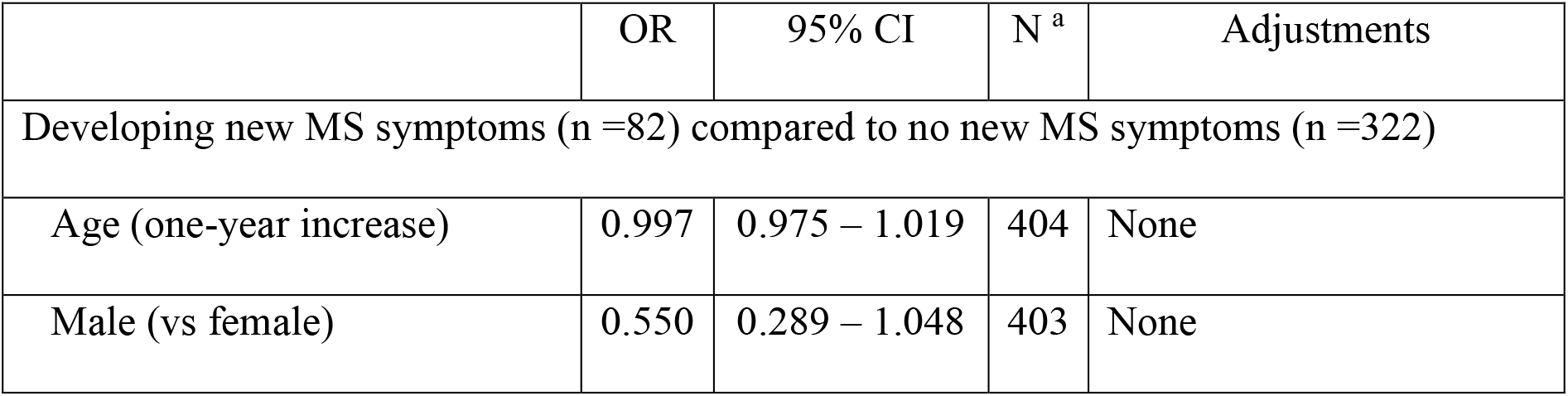
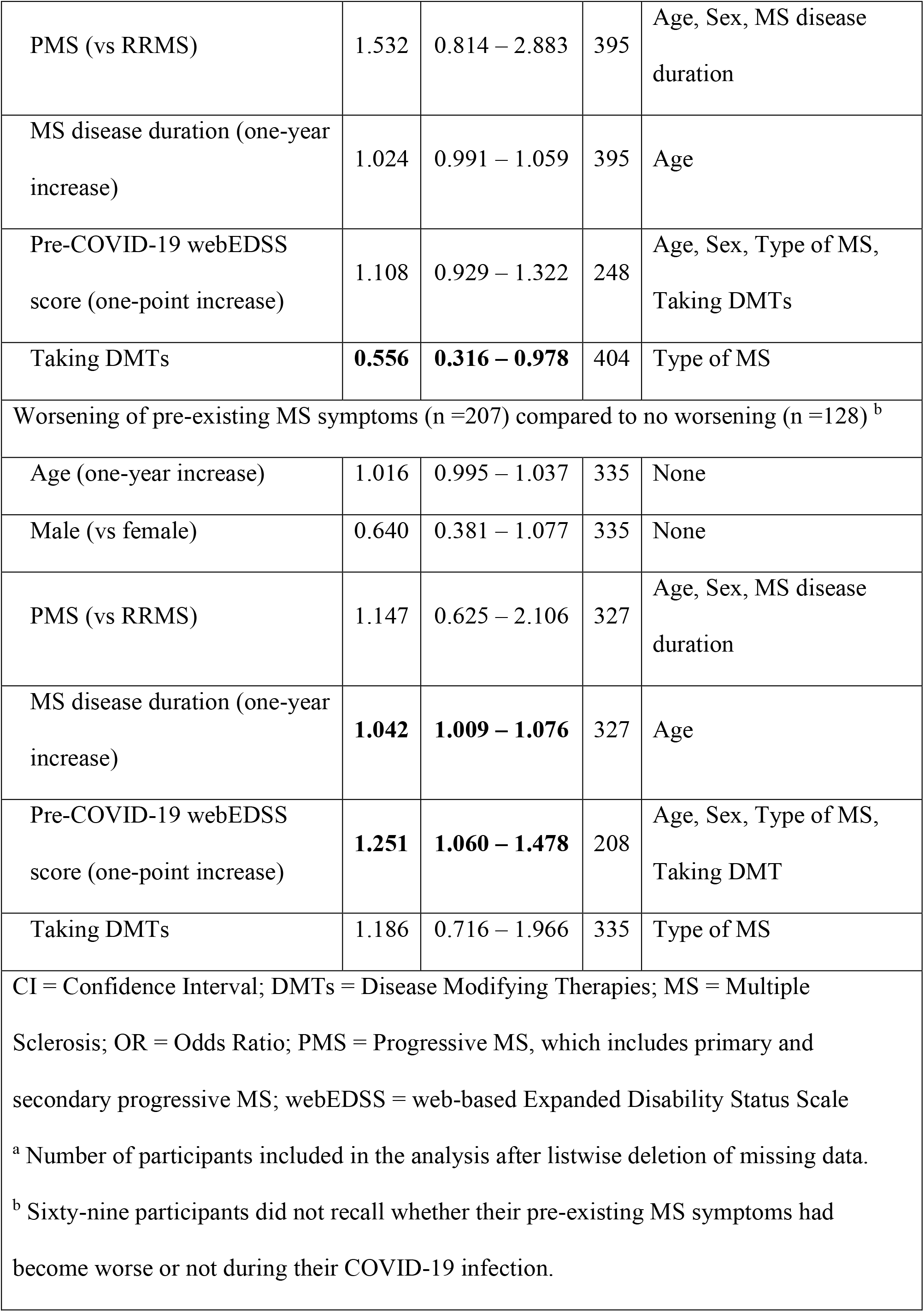
Factors associated with changes in symptoms of multiple sclerosis

|  | OR | 95% CI | N <sup>a</sup> | Adjustments |
| --- | --- | --- | --- | --- |
| Developing new MS symptoms (n =82) compared to no new MS symptoms (n =322) |  |  |  |  |
| Age (one-year increase) | 0.997 | 0.975 – 1.019 | 404 | None |
| Male (vs female) | 0.550 | 0.289 – 1.048 | 403 | None |
| PMS (vs RRMS) | 1.532 | 0.814 – 2.883 | 395 | Age, Sex, MS disease duration |
| MS disease duration (one-year increase) | 1.024 | 0.991 – 1.059 | 395 | Age |
| Pre-COVID-19 webEDSS score (one-point increase) | 1.108 | 0.929 – 1.322 | 248 | Age, Sex, Type of MS, Taking DMTs |
| Taking DMTs | **0.556** | **0.316 – 0.978** | 404 | Type of MS |
| Worsening of pre-existing MS symptoms (n =207) compared to no worsening (n =128) <sup>b</sup> |  |  |  |  |
| Age (one-year increase) | 1.016 | 0.995 – 1.037 | 335 | None |
| Male (vs female) | 0.640 | 0.381 – 1.077 | 335 | None |
| PMS (vs RRMS) | 1.147 | 0.625 – 2.106 | 327 | Age, Sex, MS disease duration |
| MS disease duration (one-year increase) | **1.042** | **1.009 – 1.076** | 327 | Age |
| Pre-COVID-19 webEDSS score (one-point increase) | **1.251** | **1.060 – 1.478** | 208 | Age, Sex, Type of MS, Taking DMT |
| Taking DMTs | 1.186 | 0.716 – 1.966 | 335 | Type of MS |
| CI = Confidence Interval; DMTs = Disease Modifying Therapies; MS = Multiple Sclerosis; OR = Odds Ratio; PMS = Progressive MS, which includes primary and secondary progressive MS; webEDSS = web-based Expanded Disability Status Scale<br><sup>a</sup> Number of participants included in the analysis after listwise deletion of missing data.<br><sup>b</sup> Sixty-nine participants did not recall whether their pre-existing MS symptoms had become worse or not during their COVID-19 infection. |  |  |  |  |

**Table 4.** Characteristics of participants with and without new symptoms of multiple sclerosis during COVID-19 infection

|  | With<br>new MS symptoms<br>n =82 | Without<br>new MS symptoms<br>n =322 | *p* value |
| --- | --- | --- | --- |
| Age, years, mean (SD) | 50 (11) | 50 (11) | 0.784 |
| Female, n (%) | 68 (82.9) | 239 (74.2) | 0.066 |
| White ethnicity, n (%) | 3 (3.7) | 21 (6.5) | 0.327 |
| Pre-COVID-19 webEDSS<br>score, median (IQR) | 5 (2.875 – 6.5)<br>n =50 | 4 (3 – 6.5)<br>n =198 | 0.481 |
| MS type, n (%) |  |  |  |
| RRMS | 53 (64.6) | 224 (69.6) | 0.589 <sup>a</sup> |
| SPMS | 14 (17.1) | 51 (15.8) |  |
| PPMS | 11 (13.4) | 28 (8.7) |  |
| Unknown | 4 (4.9) | 19 (5.9) |  |
| MS disease duration, years,<br>median (IQR) | 11.5 (5 – 20.5)<br>n =80 | 11 (6 – 17)<br>n =315 | 0.564 |
| DMTs <sup>b</sup>, n (%) | 30 (36.6) | 163 (50.6) | 0.023 |
| Beta interferons | 4 (13.3) | 17 (10.4) |  |
| Glatiramer acetate | 6 (20) | 16 (9.8) |  |
| Teriflunomide | 2 (6.7) | 5 (3.1) |  |
| Dimethyl fumarate | 8 (26.7) | 50 (30.7) |  |
| Fingolimod | 2 (6.7) | 22 (13.5) |  |
| Natalizumab | 5 (16.7) | 19 (11.7) |  |
| Ocrelizumab | 1 (3.3) | 13 (8) |  |
| Cladribine | 0 (0) | 7 (4.3) |  |
| Alemtuzumab | 2 (6.7) | 11 (6.7) |  |
| Others <sup>c</sup> | 0 (0) | 3 (1.8) |  |
| Required more help <sup>d</sup>, n (%) | 28 (39.4) | 68 (23.9) | 0.009 |

DMTs = Disease Modifying Therapies; IQR = Interquartile Range; PPMS = Primary Progressive MS; RRMS = Relapsing Remitting MS; SPMS = Secondary Progressive MS; webEDSS = web-based Expanded Disability Status Scale
<sup>a</sup> One cell (12.5%) has expected count less than 5.
<sup>b</sup> Percentages of individual DMTs are calculated based on the total number of participants taking DMTs in each group.
<sup>c</sup> Participants were taking Ponesimod (n =1) and Rituximab (n =2).
<sup>d</sup> During their COVID-19 infection than before.

Thirty-six (44%) participants with new MS symptoms reported recovery from these symptoms; 21 (26%) recovered within three weeks. Among the 46 participants who had not reported recovery, the median (IQR) duration from reporting COVID-19 to reporting persistence of the symptoms was 14 (10 – 17) weeks.

### Pre-existing MS Symptoms

Among the 207 participants with worsened pre-existing MS symptoms during the infection (Table 5), 190 (92%) reported this worsening to be the same as (n=91) or worse than (n=99) their previous non-COVID-19 systemic infection.

**Table 5.** Characteristics of participants with and without worsened pre-existing symptoms of multiple sclerosis during COVID-19 infection

|  | With worsened<br>pre-existing MS<br>symptoms<br>n =207 | Without worsened<br>pre-existing MS<br>symptoms<br>n =128 <sup>a</sup> | *p* value |
| --- | --- | --- | --- |
| Age, years, mean (SD) | 51 (11) | 49 (11) | 0.140 |
| Female, n (%) | 166 (80.2) | 93 (72.7) | 0.176 |
| White ethnicity, n (%) | 197 (95.2) | 117 (91.4) | 0.167 |
| Pre-COVID-19 webEDSS<br>score, median (IQR) | 4.5 (3 – 6.5)<br>n =133 | 4 (2.5 – 6.5)<br>n =75 | 0.035 |
| MS type, n (%) |  |  |  |
| RRMS | 138 (66.7) | 90 (70.3) | 0.648 |
| SPMS | 36 (17.4) | 18 (14.1) |  |
| PPMS | 21 (10.1) | 10 (7.8) |  |
| Unknown | 12 (5.8) | 10 (7.8) |  |
| MS disease duration, years,<br>median (IQR) | 12 (7 – 19)<br>n =203 | 8 (4 – 15.75)<br>n =124 | 0.001 |
| DMTs <sup>b</sup>, n (%) | 101 (48.8) | 61 (47.7) | 0.840 |
| Beta interferons | 9 (8.9) | 9 (14.8) |  |
| Glatiramer acetate | 11 (10.9) | 6 (9.8) |  |
| Teriflunomide | 3 (3) | 1 (1.6) |  |
| Dimethyl fumarate | 36 (35.6) | 14 (23) |  |
| Fingolimod | 13 (12.9) | 9 (14.8) |  |
| Natalizumab | 13 (12.9) | 8 (13.1) |  |
| Ocrelizumab | 6 (5.9) | 5 (8.2) |  |
| Cladribine | 3 (3) | 4 (6.6) |  |
| Alemtuzumab | 5 (5) | 5 (8.2) |  |
| Others <sup>c</sup> | 2 (1.2) | 0 (0) |  |
| Required more help <sup>d</sup>, n (%) | 70 (39.8) | 8 (6.6) | <0.001 |

DMTs = Disease Modifying Therapies; IQR = Interquartile Range; PPMS = Primary Progressive MS; RRMS = Relapsing Remitting MS; SPMS = Secondary Progressive MS; webEDSS = web-based Expanded Disability Status Scale
<sup>a</sup> Sixty-nine participants did not recall whether their pre-existing MS symptoms had become worse or not during their COVID-19 infection.
<sup>b</sup> Percentages of individual DMTs are calculated based on the total number of participants taking DMTs in each group.
<sup>c</sup> Participants were taking Ponesimod (n =1) and Rituximab (n =1).
<sup>d</sup> During their COVID-19 infection than before.

The pre-existing MS symptoms of participants with a higher pre-COVID-19 webEDSS score (adjusted OR 1.251, 95% CI 1.060 – 1.478) and longer MS disease duration (adjusted OR 1.042, 95% CI 1.009 – 1.076) were more likely to worsen during the infection (Table 3).

Sixty-three (30%) participants who experienced worsening of their pre-existing MS symptoms during the infection reported returning to baseline; 42 (20%) recovered within three weeks. Among the 144 participants who had not returned to baseline, the median (IQR) duration from reporting COVID-19 to responding to the questionnaire was 14 (9 – 16) weeks.

## DISCUSSION

This large community-based study found that 57% of people with MS and COVID-19 experience an MS exacerbation during their infection, including 20% who develop new MS symptoms. Previous studies have demonstrated an increased risk of MS exacerbations associated with other infections,^1^ but the rates (9% to 41%) ^10-14^ are lower than COVID-19 related exacerbations reported in this study. We could not objectively assess the reported new MS symptoms by neurological examination to confirm that they were relapses due to the restrictions caused by the pandemic. Previously, it has been shown that relapses reported by people with MS are often also diagnosed as relapses by clinicians ^15^.

An association between DMT use and reduction of infection-related exacerbations of MS has not been conclusively established ^11 13^. We found that taking a DMT reduces the probability of developing new MS symptoms during COVID-19 infection by 44%, which is consistent with the overall relapse rate reduction observed in clinical trials of current DMTs ^16^. Our data suggest that different DMTs might have a variable effect in preventing COVID-19 related new MS symptoms. This very preliminary finding is interesting but needs to be confirmed in larger studies.

Studies have suggested that infection-related exacerbations can be more severe and prolonged compared to exacerbations not induced by an infection ^13 14^. In our study, the MS exacerbation of many participants had not resolved three months after their COVID-19 infection. Most individuals with COVID-19 related worsening of their MS symptoms reported a deterioration that was worse than or similar to their previous non-COVID-19 infection. This finding could have been influenced by recall bias, however. In addition, most individuals reported that their new MS symptoms resulted in limitation of their daily activities.

We wondered whether people had regarded their COVID-19 symptoms, such as fatigue or cognitive problems that can mimic MS symptoms, as deterioration of their MS. Can we truly distinguish MS deterioration from some systemic symptoms of COVID-19? We cannot answer this question with confidence without paraclinical tests, but we found that most individuals with fatigue, memory, or mobility problems also reported other neurological symptoms suggestive of MS.

Although more individuals with anxiety or depression reported an MS exacerbation during their COVID-19 infection than individuals without anxiety or depression, the rate of MS exacerbations was above 50% in both groups, suggesting that over-reporting of symptoms linked to anxiety or depression has not driven these results ^17^.

## CONCLUSIONS

In this study, we demonstrate that COVID-19 is associated with MS exacerbations. This finding highlights the importance of protecting people with MS against the infection which is now feasible with the increasing number of COVID-19 vaccines. Fewer people taking DMTs experience new neurological symptoms following COVID-19, and, therefore, it is important to consider carefully before altering or delaying treatment with DMTs because of concerns about their safety during the pandemic.

## Supporting information

STROBE

